# Variation in APOE ε4 Prevalence and Brain Health Associations by European Descent in White All of Us Participants

**DOI:** 10.1101/2025.07.16.25331679

**Authors:** Jaime Perales-Puchalt, Clayton O. Mansel, Olivia J. Veatch

## Abstract

**Importance:** The *APOE* ε4 allele is among the best-established genetic risk factor for Alzheimer’s disease. Different ethnic and racial groups have varying *APOE* ε4 prevalences. The prevalence of *APOE* ε4 also varies among European regions, yet White Americans are treated as a monolith, potentially impacting recruitment decisions and risk profile estimations.

**Objective:** To test whether White Americans of Southern European descent have a lower prevalence of *APOE* ε4 than their Northern European descent peers, and secondarily, if *APOE* ε4-brain health outcomes vary by European descent.

**Design, setting and participants:** Cross-sectional analyses of the nationwide All of Us cohort study in the United States. Overall inclusion criteria were reporting only White race, and countries of descent from Northern, Central or Southern Europe without overlap. Enrollment opened in May 2018. Analyses were conducted April-June 2025.

**Exposures:** Regions of European descent (Northern, Central and Southern) from the Basics Survey.

**Main Outcomes and Measures:** APOE ε4 using variants rs429358 and rs7412, coded as zero, one or two copies.

**Results:** Among those aged ≥18 (n=77,676), the percentage of European descent was 7.7% Southern and 64.7% Northern. Age and female sex at birth were 55.5 years and 55.8% among Americans of Southern and 56.3 years and 57.4% among those of Northern European descent. APOE ε4 allele frequency was 17.4% for one and 0.1% for two copies among Americans of Southern, and 24.9% and 2.0% for Northern European descent. Logistic regression models confirmed these differences: compared to Americans of Southern European descent, those of Northern European descent had an Odds Ratio of having one APOE ε4 allele of 1.60 (1.49-1.71), and 2.21 (1.71-2.88) of having two APOE ε4 alleles. European descent moderated associations between APOE ε4 allele frequency and two brain health outcomes (interaction p≤0.10).

**Conclusions and Relevance:** The frequency of *APOE* ε4 among Americans of Southern European descent was lower than that of Northern European descent and associations with some brain health outcomes varied. Future studies need to use probabilistic samples to confirm these findings. The concept of White race might be too broad when making racial comparisons of *APOE* ε4 frequency and its effects.

**Key Points:** *Question:* Is the prevalence of *APOE* ε4 reduced when comparing Southern European to Northern European descent American adults who identify as only White race?

*Findings:* In this cross-sectional analysis of a cohort study (n=77,676 American adults), those of Northern European descent had 60% higher odds of having one, and 120% of having two copies of *APOE* ε4 compared to those of Southern European descent.

*Meaning:* Among White American adults, the prevalence of *APOE* ε4 differs by region of European descent; future studies need to confirm these findings using genetically inferred ancestry and probabilistic samples with stronger external validity.

## Introduction

Apolipoprotein ε4 (*APOE* ε4) is the strongest allelic risk factor for late-onset Alzheimer’s disease (AD).^1^ US-based studies suggest that the prevalence and effects of *APOE* ε4 vary by race/ethnicity.^2^ These ethnoracial differences matter because *APOE* ε4 is often related to adverse events and outcomes,^3,4^ and self-identified ethnoracial data provide an inexpensive approach to make recruitment decisions, estimating risk profiles, and tailoring interventions. However, ethnoracial classifications ignore within-group variability.

White Americans are largely of European descent,^5^ and are the largest group in the US population and dementia trials.^6^ Previous studies in Europe show that *APOE* ε4 prevalence is lower among Southern, than among Northen Europeans with AD.^7,8^ Europeans and European Americans from these regions can be differentiated consistently in a genome-wide single nucleotide polymorphism panel.^9^ In this manuscript, we tested whether the prevalence of *APOE* ε4 and its association with brain health outcomes differ between Americans of Northern (AmEuro-N) and Southern European descent (AmEuro-S). In line with previous research,^7,8^ we hypothesize that White AmEuro-S will have a lower prevalence of *APOE* ε4 than AmEuro-N.

## Methods

### Sample

This was a cross-sectional secondary analysis of the All of Us cohort study.^10^ This study opened for enrollment in May 2018 and consists of a convenience sample of participants ≥18 years old who were recruited via multiple approaches at more than 340 recruitment sites across the US. The study collects health questionnaires, electronic health records (EHRs), and biospecimens. Informed consent is conducted in person or through eConsent. The protocol was reviewed by the Institutional Review Board (IRB) of the All of Us Research Program.

We selected three subsamples of participants:

- Subsample 1 (APOE ε4 prevalence): Age at consent ≥18; available *APOE* ε4 genotypes; White race only; descend from Northern, Central or Southern European countries without overlap.
- Subsample 2a and 2b (health survey *APOE* ε4-brain health association): Same as ‘Subsample 1’ plus age ≥60; no missing data on sex at birth and health survey outcomes.
- Subsample 3 (EHRs *APOE* ε4-brain health association): Same as ‘Subsample 2’ but no missing EHR-derived outcome data instead of health survey outcomes.

### Race, Ethnicity, Country of Descent Definitions, and APOE Genotyping

Self-reported Race, Ethnicity and country/es of descent were drawn from the Basics survey. We categorized European descent into Northern, Central, and Southern based on previous studies.^7,8^ *APOE* ε4 genotypes (primary outcome) were ascertained from single-nucleotide variants from the All of Us Allele Count/Allele Frequency unphased callset (rs429358 and rs7412), which encompasses variants exceeding a population-specific minor allele frequency threshold of >1% or allele count depth >100.^11^ Genotype extraction was performed using PLINK 1.9.12 and imputed with R as previously described.^11^

### Secondary Brain Health Outcomes

The first outcome, from the Behavioral Health and Personality survey, was self-reported frequency of memory problems based on the question, “How often do you have problems remembering appointments or obligations?” Responses (five-point Likert scale) were dichotomized into never vs sometimes to very often. The second outcome, from the Personal and Family Health survey, was self-reported history of dementia or cognitive impairment (yes/no), based on whether participants endorsed themselves in response to either of two questions about family history of dementia or memory loss. The third outcome was dementia diagnosis (yes/no), ascertained from EHRs and defined using a validated computable phenotype with excellent predictive value.^13^ Participants were labelled as a dementia case if they had five or more visits with matching ICD-9/ICD-10 codes or one matching drug prescription.

### Statistical Analysis

We conducted analyses between April and June 2025. We used descriptive statistics to characterize the subsamples. The association between *APOE* genotype and European country descent group was analyzed using a multinomial logistic regression with Southern European set as the reference group using the *nnet* package in R. In Subsample 1, models were analyzed raw and controlling for ethnicity, as suggested by literature.^2^ The results are presented as odds ratios (95% confidence interval) and p values were calculated using Wald tests. For subsamples 2a, 2b, and 3, the associations between *APOE* genotype (0 vs 1 *APOE* ε4 copy and 0 vs 2 copies) and the outcome were analyzed using multivariable logistic regression models stratified by European descent country group. Models were adjusted for ethnicity, sex assigned at birth, and age, as suggested by literature.^2^ We used Likelihood Ratio tests comparing the additive to the multiplicative model to test for interactions between *APOE* genotype and European country descent group. The threshold for statistical significance for all non-interactive analyses was set at α=0.05, as we only proposed one hypothesis and all other tests were exploratory. For the interaction tests, the threshold was set at α=0.10. All analyses were conducted using R version 4.4.0 and RStudio version 2024.04.0 within the All of Us Researcher Workbench.

## Results

Out of a total of 133,074 participants who reported only White race, 77,676 participants were included in subsample 1, 10,112 in subsample 2a, 24,274 in subsample 2b, and 39,197 in subsample 3 (**Table 1**). In Subsample 1, the most frequent countries were England (67.7%) and Ireland (37.0%; AmEuro-N), Germany (70.8%) and Poland (33.3%; Central: AmEuro-C), and Italy (86.6%) and Spain (16.1%; AmEuro-S). APOE ε4 allele prevalence was 17.4% for one and 0.1% for two copies among AmEuro-S, 22.5% and 1.7% for AmEuro-C and 24.9% and 2.0% for AmEuro-N. Logistic regressions showed that AmEuro-C and AmEuro-N had a significantly higher *APOE* ε4 prevalence thano AmEuro-S (**Table 2**). Odds ratios increased slightly when including only those ≥60 years old and barely changed when controlling for Latino ethnicity.

**Table 1.**
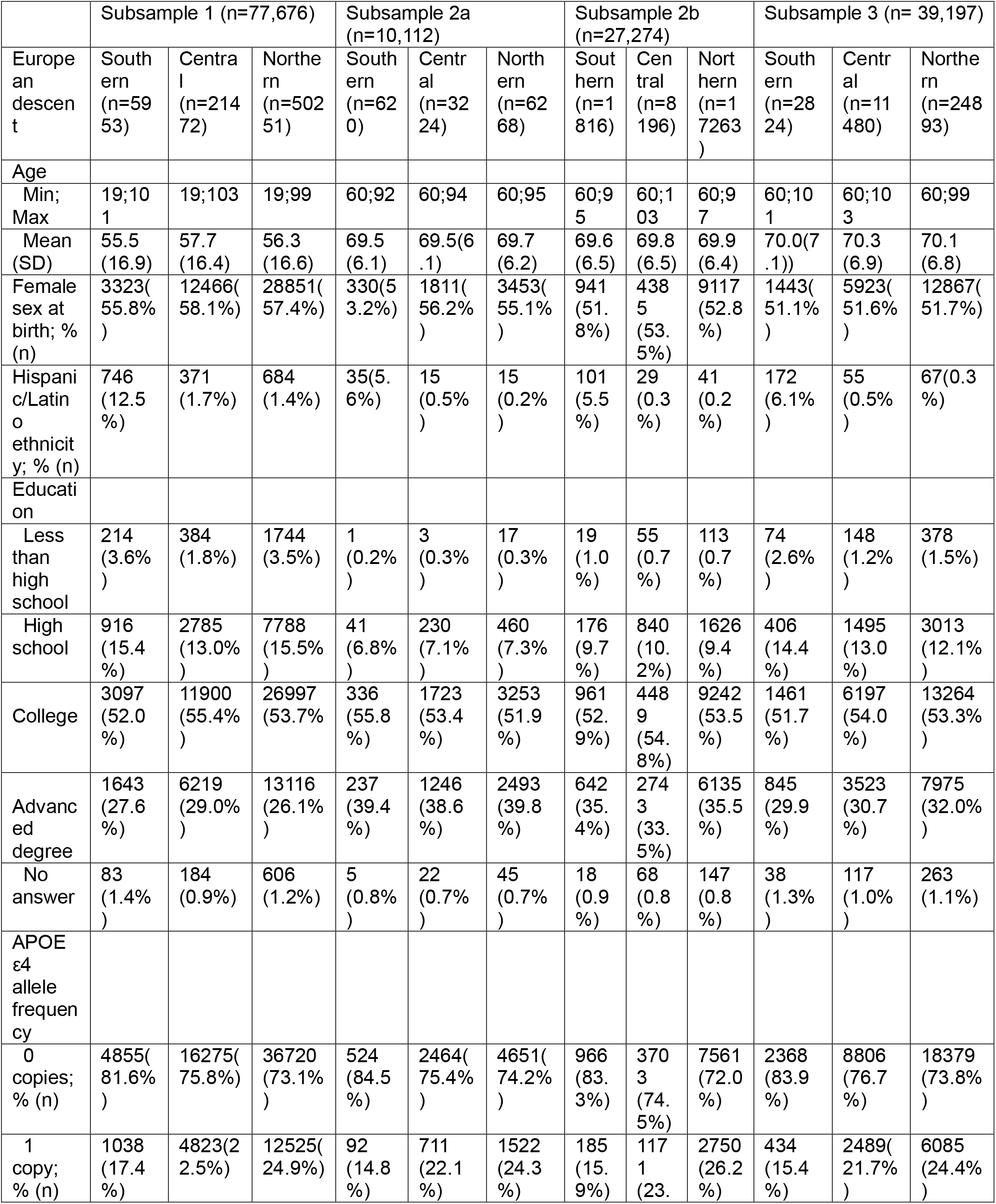
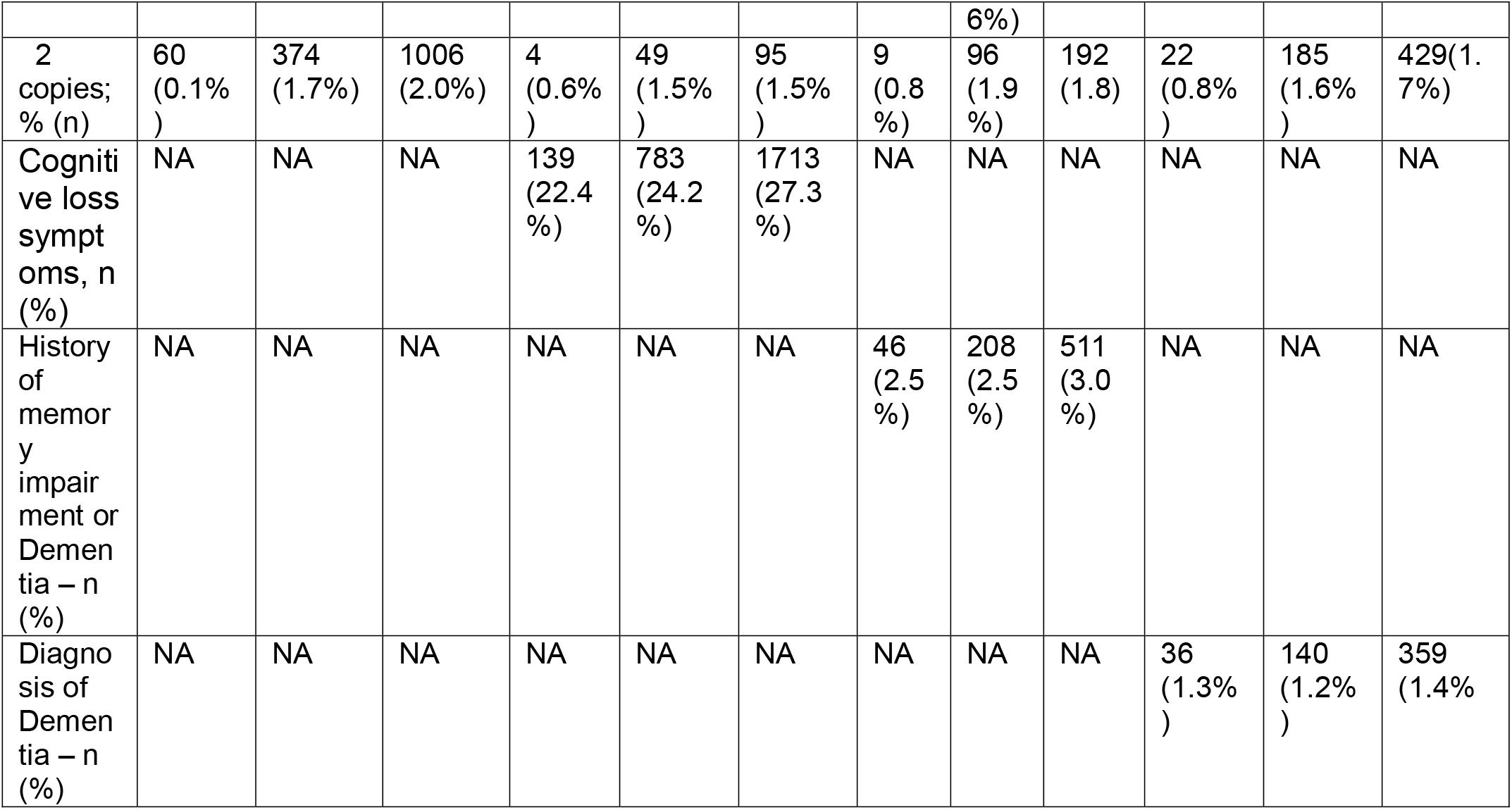
Characteristics of the subsamples.

**Table 2.** Associations between descent groups and APOE ε4 status in subsample 1.

|  | Among those 18 and older<br>(n=77,676) |  | Among those 60 and older<br>(n=39,197) |  |
| --- | --- | --- | --- | --- |
|  | OR (95% CI); p value |  | OR (95% CI); p value |  |
|  | E4/- | E4/E4 | E4/- | E4/E4 |
| European descent (raw) |  |  |  |  |
| Southern | Ref | Ref | Ref | Ref |
| Central | **1.38 (1.29;1.49);**<br>**<0.001** | **1.86 (1.41;2.45);**<br>**<0.001** | **1.54 (1.38;1.72);**<br>**<0.001** | **2.26 (1.45;3.52);**<br>**<0.001** |
| Northern | **1.60 (1.49;1.71);**<br>**<0.001** | **2.21(1.71;2.88);**<br>**<0.001** | **1.81 (1.62;2.01);**<br>**<0.001** | **2.51 (1.63;3.86);**<br>**<0.001** |
| European descent (adjusted for Latino ethnicity) |  |  |  |  |
| Southern | Ref | Ref | Ref | Ref |
| Central | **1.39 (1.29;1.50);**<br>**<0.001** | **1.77 (1.34;2.33);**<br>**<0.001** | **1.55 (1.39;1.73);**<br>**<0.001** | **2.13 (1.37;3.32);**<br>**<0.001** |
| Northern | **1.60 (1.49;1.72);**<br>**<0.001** | **2.11 (1.62;2.75);**<br>**<0.001** | **1.82 (1.63;2.02);**<br>**<0.001** | **2.36 (1.53;3.64);**<br>**<0.001** |

**Bold** indicate significance (p<0.05)

Associations between *APOE* ε4 frequency and self-reported memory loss and dementia diagnosis were modified by descent (interaction p≤0.10; **Table 3**). Trends generally showed stronger associations between *APOE* ε4 and outcomes among AmEuro-N than AmEuro-S, and were statistically significant among AmEuro-N but not AmEuro-S. An outlier was the association between two vs zero copies of *APOE* ε4 on dementia diagnosis, which was stronger among AmEuro-S than AmEuro-N (p<0.05).

**Table 3.** Associations between APOE ε4 status and brain health outcomes by descent group.

| Subsample 2a |  |  |  |  |  |  |  |
| --- | --- | --- | --- | --- | --- | --- | --- |
|  | Among Southern European descent (n=620) |  | Among Central European descent (n=3,224) |  | Among Northern European descent (n=6,268) |  | p of interaction between APOE ε4 and European descent |
| APOE ε4 on | OR | (95%CI); p value | OR | (95%CI); p value | OR | (95%CI); p value |  |
| Cognitive loss symptoms (raw) |  |  |  |  |  |  |  |
| One copy vs 0 (Ref) | 1.02 | (0.59;1.70);0.94 | 1.00 | (0.82;1.21);0.98 | 1.11 | (0.98;1.26);0.11 | 0.49 |
| Two copies vs 0 (Ref) | NA | NA | 1.25 | (0.65;2.29);0.48 | 1.33 | (0.85;2.03);0.19 |  |
| Cognitive loss symptoms (adjusted for ethnicity, sex at birth and age) |  |  |  |  |  |  |  |
| One copy vs 0 (Ref) | 1.01 | (0.58;1.70);0.96 | 1.00 | (0.82;1.22);0.96 | 1.11 | (0.97;1.26);0.12 | 0.51 |
| Two copies vs 0 (Ref) | NA | NA | 1.32 | (0.68-2.44) | 1.32 | (0.84;2.02);0.21 |  |
| Subsample 2b |  |  |  |  |  |  |  |
|  | Among Southern European descent (n=1816) |  | Among Central European descent (n=8195) |  | Among Northern European descent (n=17263) |  |  |
| APOE ε4 on | OR | (95%CI); p value | OR | (95%CI); p value | OR | (95%CI); p value |  |
| Self-reported memory loss or AD (raw) |  |  |  |  |  |  |  |
| One copy vs 0 (Ref) | 1.18 | (0.51;2.43);0.67 | 0.88 | (0.61;1.23);0.45 | **1.43** | **(1.17;1.73);<0.001** | **0.09** |
| Two copies vs 0 (Ref) | N/A | N/A | 1.49 | (0.52;3.34);0.38 | **2.62** | **(1.57;4.11);<0.001** |  |
| Self-reported memory loss or AD (adjusted for ethnicity, sex at birth and age) |  |  |  |  |  |  |  |
| One copy vs 0 (Ref) | 1.20 | (0.51-2.47);0.65 | **1.52** | **(1.32;1.73);<0.001** | **1.43** | **(1.18;1.74);<0.001** | **0.10** |
| Two copies | N/A | N/A | **1.82** | **(1.21;2.74);0.004** | **2.62** | **(1.57;4.12);<0.001** |  |
| vs 0 (Ref) |  |  |  |  |  |  |  |
| Subsample 3 |  |  |  |  |  |  |  |
|  | Among Southern European descent (n=2,824) |  | Among Central European descent (n=11,480) |  | Among Northern European descent (n=24,893) |  |  |
| APOE ε4 on | OR | (95%CI); p value | OR | (95%CI); p value | OR | (95%CI); p value |  |
| Dementia diagnosis (raw) |  |  |  |  |  |  |  |
| One copy vs 0 (Ref) | 1.42 | (0.57;3.11);0.41 | 1.37 | (0.91;2.00);0.12 | **1.74** | **(1.39;2.17);<0.001** | **0.07** |
| Two copies vs 0 (Ref) | **8.67** | **(1.34;31.78);0.004** | **7.84** | **(4.20;13.60);<0.001** | **3.00** | **(1.69;4.94);<0.001** |  |
| Dementia diagnosis (adjusted for ethnicity, sex at birth and age) |  |  |  |  |  |  |  |
| One copy vs 0 (Ref) | 1.49 | (0.59;3.27);0.35 | 1.42 | (0.94;2.08);0.08 | **1.84** | **(1.47;2.29);<0.001** | **0.08** |
| Two copies vs 0 (Ref) | **9.72** | **(1.49;36.66);0.003** | **8.63** | **(4.60;15.08);<0.001** | **3.45** | **(1.93;5.71);<0.001** |  |

**Bold** indicate significance (p<0.05 or Wald 95% confidence limits)
N/A indicates that there were no individuals with that APOE genotype in the subsample

## Discussion

To our knowledge, this is the first study to explore the variation in APOE ε4 prevalence and brain health associations by European descent in White US individuals. In line with our hypothesis, we showed that the prevalence of *APOE* ε4 among AmEuro-S was lower than that of AmEuro-N. Like previous research,^7,8^ prevalence among AmEuro-C resembled that of AmEuro-N. European descent moderated associations between *APOE* ε4 frequency and some brain health outcomes.

Study limitations included 1) a non-population-based sample, with questionable generalizability, despite prevalence estimates that resemble European population-based samples (e.g., 14.4% in Spain and 27.0% in the UK);^7,8^ 2) lower sample sizes among AmEuro-S; 3) the use of EHR billing codes instead of clinical protocols to identify dementia cases, and 4) not accounting for intra-country descent variability.

Future studies should 1) use probabilistic samples to confirm these findings, 2) assess AD and cognitive impairment using standard criteria, and explore 3) operationalizations to represent Americans of mixed European descent, 4) effect mechanisms, and 5) the role of genetic ancestry.

We conclude that the concept of White race might be too broad when comparing *APOE* ε4 prevalence and risk among races and ethnicities. Researchers need to consider intra-group European descent differences among White Americans for power-sampling, recruitment, developing risk profiles and plan for adverse effects.

## Data Sharing Statement

### Data

### Data Available

No

### Additional Information

Data from the All of Us Research Program are publicly available at www.allofus.nih.gov. We used release version 8, which includes participant data with a cutoff date of October 1, 2023. All data access and analyses were conducted within a secure informatic workspace provided by the National Institutes of Health.

## Data Availability statement

All data used in this study can be accessed by qualified researchers at allofus.nih.gov. Source code for these analyses is published as a Community Workspace within the *All of Us* Researcher Workbench and is available at (https://workbench.researchallofus.org/workspaces/aou-rw-662ce24a/europeandescentofwhiteamericansandapolipoprotein4/data)

## Acknowledgements

We acknowledge *All of Us* participants for their contributions, without whom this research would not have been possible. We also thank the National Institutes of Health’s *All of Us* Research Program for making available the participant and genetic data examined in this study. The All of Us Research Program is supported by the National Institutes of Health, Office of the Director: Regional Medical Centers: 1 OT2 OD026549; 1 OT2 OD026554; 1 OT2 OD026557; 1 OT2 OD026556; 1 OT2 OD026550; 1 OT2 OD 026552; 1 OT2 OD026553; 1 OT2 OD026548; 1OT2 OD026551; 1 OT2 OD026555; IAA #: AOD 16037; Federally Qualified Health Centers: HHSN 263201600085U; Data and Research Center: 5 U2C OD023196; Biobank: 1 U24 OD023121; The Participant Center: U24 OD023176; Participant Technology Systems Center: 1 U24 OD023163; Communications and Engagement: 3 OT2 OD023205; 3 OT2 OD023206; and Community Partners: 1 OT2 OD025277; 3 OT2 OD025315; 1 OT2 OD025337; 1 OT2 OD025276. In addition, the *All of Us* Research Program would not be possible without the partnership of its participants.

## References

1. Long JM, Holtzman DM. Alzheimer Disease: An Update on Pathobiology and Treatment Strategies. Cell. 2019;179(2):312–339. doi:10.1016/j.cell.2019.09.001

2. Jackson RJ, Hyman BT, Serrano-Pozo A. Multifaceted roles of APOE in Alzheimer disease. Nat Rev Neurol. Aug 2024;20(8):457–474. doi:10.1038/s41582-024-00988-2

3. Budd Haeberlein S, Aisen P, Barkhof F, et al. Two randomized phase 3 studies of aducanumab in early Alzheimer’s disease. The journal of prevention of Alzheimer’s disease. 2022;9(2):197–210.

4. Evans CD, Sparks J, Andersen SW, et al. APOE ε4’s impact on response to amyloid therapies in early symptomatic Alzheimer’s disease: Analyses from multiple clinical trials. Alzheimer’s & Dementia. 2023;19(12):5407–5417.

5. US Census Bureau. About the topic of race. https://www.census.gov/topics/population/race/about.html#:∼:text=White%20%E2%80%93%20A%20person%20having%20origins,Black%20racial%20groups%20of%20Africa.

6. Faison WE, Schultz SK, Aerssens J, et al. Potential ethnic modifiers in the assessment and treatment of Alzheimer’s disease: challenges for the future. International psychogeriatrics. 2007;19(3):539–558. doi:10.1017/S104161020700511X

7. Ward A, Crean S, Mercaldi CJ, et al. Prevalence of apolipoprotein E4 genotype and homozygotes (APOE e4/4) among patients diagnosed with Alzheimer’s disease: a systematic review and meta-analysis. Neuroepidemiology. 2012;38(1):1–17. doi:10.1159/000334607

8. Crean S, Ward A, Mercaldi CJ, et al. Apolipoprotein E ε4 prevalence in Alzheimer’s disease patients varies across global populations: a systematic literature review and meta-analysis. Dement Geriatr Cogn Disord. 2011;31(1):20–30. doi:10.1159/000321984

9. Seldin MF, Shigeta R, Villoslada P, et al. European population substructure: clustering of northern and southern populations. PLoS Genet. Sep 15 2006;2(9):e143. doi:10.1371/journal.pgen.0020143

10. The “All of Us” Research Program. New England Journal of Medicine. 2019;381(7):668–676. doi:doi:10.1056/NEJMsr1809937

11. Khajouei E, Ghisays V, Piras IS, et al. Phenome-Wide Association of APOE Alleles in the All of Us Research Program. medRxiv. Sep 4 2024;doi:10.1101/2024.09.04.24313010

12. Chang CC, Chow CC, Tellier LC, Vattikuti S, Purcell SM, Lee JJ. Second-generation PLINK: rising to the challenge of larger and richer datasets. Gigascience. 2015;4:7. doi:10.1186/s13742-015-0047-8

13. Ritchie MD, Denny JC, Crawford DC, et al. Robust replication of genotype-phenotype associations across multiple diseases in an electronic medical record. The American Journal of Human Genetics. 2010;86(4):560–572.

